# Unravelling the Impact of Tumor Location on Patient Survival in Glioblastoma: A Genomics and Radiomics Approach

**DOI:** 10.1101/2024.10.02.24314823

**Authors:** Kavita Kundal, Neeraj Kumar, Rahul Kumar

## Abstract

Glioblastoma (GBM), the most aggressive form of brain tumor, has a median survival rate of 12-15 months. Understanding the relationship between genetics and tumor location, as well as identifying non-invasive biomarkers, is crucial for improving treatment strategies and survival outcomes in GBM. In this study, we investigated the impact of tumor location on survival outcome of GBM patients along with genetic factors that influence tumor behaviour in different brain regions. Interestingly, we found that patients with parietal lobe tumors had significantly poor survival outcome compared to those with tumors in other brain regions, particularly the frontal lobe. In a comprehensive genomic analysis, we identified genetic factors, seemingly contributing to the poor survival outcomes in parietal lobe patients. We found the enrichment of *PTEN* loss-of-function mutations in parietal lobe tumors and interestingly these mutations are known to be associated with chemoresistance and poor patient survival. We also found two fusion genes i.e., *FGFR3-TACC3* and *EGFR-SEPT14*, exclusively in parietal lobe tumors, which are known to play crucial roles in tumorigenesis. Differential gene expression analysis revealed the upregulation of genes like *PITX2, HOXB13*, and *DTHD1*, which could be responsible for tumor progression in parietal lobe tumors. Conversely, the downregulation of *ALOX15* increased relapse risk. Copy number alterations, such as deletions in tumor suppressor gene (*LINC00290)*, were linked to the aggressive nature of parietal lobe tumors. Radiomic analysis revealed two key features, lower *LLL_GLDM_DependanceEntropy* and higher *HLL_firstorder_Mean*, both of which show a significant correlation with increased risk and poorer survival outcome. These findings suggest the potential for targeted therapies and personalized treatments based on tumor location, genetic profile, and radiomic markers. We anticipate that as the size of the datasets will increase for radiogenomics based studies, it will further strengthen these findings and our understanding of molecular drivers for GBM progression, treatment resistance and survival outcome.

## 1. Introduction

Gliomas are intrinsic tumors of the central nervous system (CNS), accounting for approximately 80% of malignant brain and CNS tumors. According to the WHO CNS5 2021 guidelines, gliomas with *IDH1-wt, EGFR* amplification, *TERT* promoter mutation, and chromosome7 gain and deletion of chromosome10 are categorized as Glioblastoma (grade IV brain tumor) [1]. Glioblastoma (GBM) is the most aggressive and undifferentiated brain tumor, with a median survival rate of 12-15 months. Its morphological diagnostic criteria include enhanced cellularity, necrosis, high mitotic activity, and microvascular proliferation (MVP) [2]. Glioblastoma recurrence is primarily due to the tendency of tumor cells to migrate to other brain tissues, despite standard treatments involving surgical resection followed by radiochemotherapy. Although significant progress has been made in understanding glioblastoma biology, it remains incurable, with no substantial therapeutic advances over the past decade. The current therapeutic approach includes micro-neurosurgical resection followed by chemoradiotherapy, aiming for gross total resection (GTR) of the tumor. GTR significantly impacts overall survival (OS), progression-free survival (PFS), and quality of life (QoL) of patients [3]. Achieving 100% tumor removal during surgery is partially feasible, as there might be a loss of functional brain areas, some areas might show more aggressive recurrence and cause high impact on patient survival.

Genomic differences significantly affect survival e.g., patients with *IDH1* mutations have better survival rates than those with *IDH1* wild type [4]. *MGMT*-methylated patients respond more favourably to therapy than *MGMT*-unmethylated patients [5]. Patients with *TERT* mutations have poorer survival than those with *TERT* wild type [6].

Anatomical tumor location also influences survival due to differences in functionality, the tumor microenvironment, and gene presence. Studies show that tumor location is a significant prognostic factor, with patients having right temporal lobe tumors showing poorer survival rates, while those with left temporal lobe tumors have better outcomes [7]. Tumors in the lateral ventricles are linked to lower survival rates [8], and left hemispheric tumors lead to a decline in Karnofsky Performance Status (KPS) and shorter progression-free survival [9]. Additionally, neural stem cells in the subventricular zone (SVZ) are associated with tumor progression and recurrence [10].

Certain genomic factors are also linked to specific locations. For instance, 1p/19q co-deletion predominantly occurs in Frontal lobe GBM [11], tumors that harbour *IDH1* mutations, exhibit proneural and/or proliferative gene expression, and do not demonstrate *PTEN* loss are more frequently found in the frontal lobe [12]. Some genomic factors in certain locations show a good response to treatment, such as *MGMT* promoter methylated tumors in the left temporal lobe being associated with a favourable response to radio-chemotherapy [12].

In recent years, radiomics has emerged as a valuable approach, utilizing imaging data to extract quantitative features from medical images such as MRI. These radiomic features provide a non-invasive insight into tumor heterogeneity and the microenvironment, enabling the identification of phenotypes that correlate with clinical outcomes. By offering early prognostic indicators, radiomics has the potential to guide more precise treatment strategies [13,14]. Despite the existence of an association between tumor location and genomic landscape in GBM, a comprehensive study is lacking. Additionally, no radiomics study based on tumor location and clinical outcomes has been reported yet. Tumor location is crucial for prognosis, and its accurate assessment, combined with genetic factors, and promising non-invasive markers can significantly impact treatment outcomes and survival.

In this study, we examined differential clinical outcomes based on tumor location, followed by an investigation of genomic alterations including somatic mutations, copy number variations (CNV), fusion genes and differential gene expression (DGE). We aimed to identify genetic aberrations, which might be contributing to poor survival. We also explored radiomic markers to predict clinical outcomes, which may serve as early non-invasive biomarkers of poor prognosis. This multi-level approach provides a deeper understanding of the genetic aberrations driving aggressive tumor behaviour and underscores the significance of imaging features as reliable non-invasive biomarkers for predicting clinical outcomes.

## 2. Methodology

### 2.1 Data Description

The genomics dataset “Glioblastoma TCGA 2013”, consisting of 577 subjects, was downloaded from cBioPortal [15]. MRI data of GBM patients was obtained from TCIA [16], which includes 262 subjects with multiple imaging modalities. Among these, 258 subjects were common between the “Glioblastoma TCGA 2013” and “TCIA-GBM” datasets, providing both genomics and MRI data. Of these, 178 subjects were *IDH1*-wildtype, and from this group, we selected 123 subjects that included four major imaging modalities (T1, T1ce, T2, and FLAIR) necessary for determining precise tumor locations.

### 2.2 Data Processing

The MRI data processing for 123 subjects involved converting raw DICOM images from four modalities (T1, T1ce, T2, and FLAIR) into NIfTI format using the “dcm2niix” Linux package [17]. Subsequently, the NIfTI images underwent a series of pre-processing steps using EnsembleUNets to address quality issues, motion artifacts, and misalignments. The pre-processing entailed four key steps where co-registration aligned all images onto a single plane specifically the T1ce, followed by bias correction that improved the MRI scan quality, normalization that enhanced image contrast, and finally, skull-stripping that removed the skull to focus on the brain area, mitigating intensity variations [18]. Following these steps, tumor segmentation was performed on the processed images using EnsembleUNets, a benchmarked tool from our previous study [18,19]. The resulting tumor segments and processed MRI scans were further utilized for volume extraction and tumor location identification.

### 2.3 Tumor Volume Extraction and Location Mapping

To extract the volume of each tumor from each brain lobe, we followed a two-step process that involved image registration and volume extraction. In the first step, the segmented images and processed T1 images were registered to the MNI152 standard space using the FSL tool’s “*flirt*” function. The processed T1 image is aligned to the MNI152 template utilizing the “*mutualinfo*” cost function, and the resulting transformation matrix is then applied to the segmented image, ensuring both images are properly aligned to the standard space. In the second step, volume extraction is conducted through a series of operations with FSL’s “*maths*” and “*stats*” functions. For this study, we extracted volume only using the Montreal Neurological Institute (MNI) atlas [20,21]. Initially, all regions from MNI atlases were extracted using the “*fslmaths*” function, involving thresholding and binarization. These extracted regions were then multiplied with the Region of Interest (ROI) using “*fslmaths*” again to generate a result image. The overlapping regions between the ROI and the atlas were isolated by applying further thresholding and binarization on the result image. Finally, the volume of these overlapping regions was calculated using the “*fslstats*” function, which outputs the volume data to a file [22]. This process ensures precise volume measurements of tumors within each brain lobe. Based on the extracted tumor volumes, samples were categorized according to their tumor presence in different brain locations.

### 2.4 Differential Survival Analysis

To determine the impact of tumor location on patient survival, we conducted a survival analysis using the “*survminer*” and “*survival*” packages in R software v4.2.1 [23], comparing survival rates among patients with tumors in various brain locations.

### 2.5 Comprehensive Genomic Analysis

Of 123 subjects with MRI available, mutation dataset, transcriptomics data, copy number alteration and fusion genes data were available for 77, 48, 100 and 46 subjects respectively. We conducted a comprehensive analysis using these datasets to investigate the genetic drivers of differential survival outcomes. This involved identifying prevalent mutations across different brain tumor locations by applying Fisher’s exact test, and visualization using “*maftools*” [24], copy number variation (CNV), fusion genes and differential gene expression (DGE) analysis using “*DESeq2*” [25].

### 2.6 Radiomic Analysis

To identify non-invasive markers associated with poor survival, we conducted a radiomic analysis focused on MRI based radiomic features. We extracted radiomic features from T1ce MRI images using the open-source Python package, *pyRadiomics* [26]. The extracted radiomic features encompass a range of quantitative characteristics e.g., texture, shape, and intensity-based measures that capture tumor heterogeneity. These features were Z-normalized and were systematically analysed to identify those specifically altered in the lobe associated with poor survival as compared to other lobes. Differential radiomic features (p-value < 0.05) were further subjected to multivariate Cox proportional hazards regression analysis [27] to assess their correlation with overall survival, enabling us to uncover potential non-invasive radiomic biomarkers that could predict survival outcomes based on MRI. This radiomic approach adds a critical dimension to our analysis, offering a non-invasive method for identifying key prognostic biomarkers associated with tumor location and survival.

## 3. Results

### 3.1 Mapping Tumor Location

Using the MNI atlas, we extracted tumor volume of 123 GBM samples from nine distinct brain regions i.e., caudate, cerebellum, frontal lobe, insula, occipital lobe, parietal lobe, putamen, temporal lobe, and Thalamus. After calculating the total tumor volume, a threshold of 25% was applied to determine tumor location [28]. Our analysis revealed that frontal, temporal and parietal lobes were most frequently involved regions with 48, 49 and 50 samples having tumor in these lobes respectively **(Figure 1A and Supplementary Table 1)**. Moderate involvement was also observed in the occipital lobe with 11 samples and the cerebellum with 3 samples. In contrast, minimal involvement was observed in the caudate and thalamus with only 1 sample. The insula and putamen regions did not have any samples exceeding the 25% volume threshold. A total of 37 subjects had tumors in multiple regions. This analysis highlights the predominant involvement of the frontal, parietal, and temporal lobes in tumor volume distribution.

**Figure 1.**
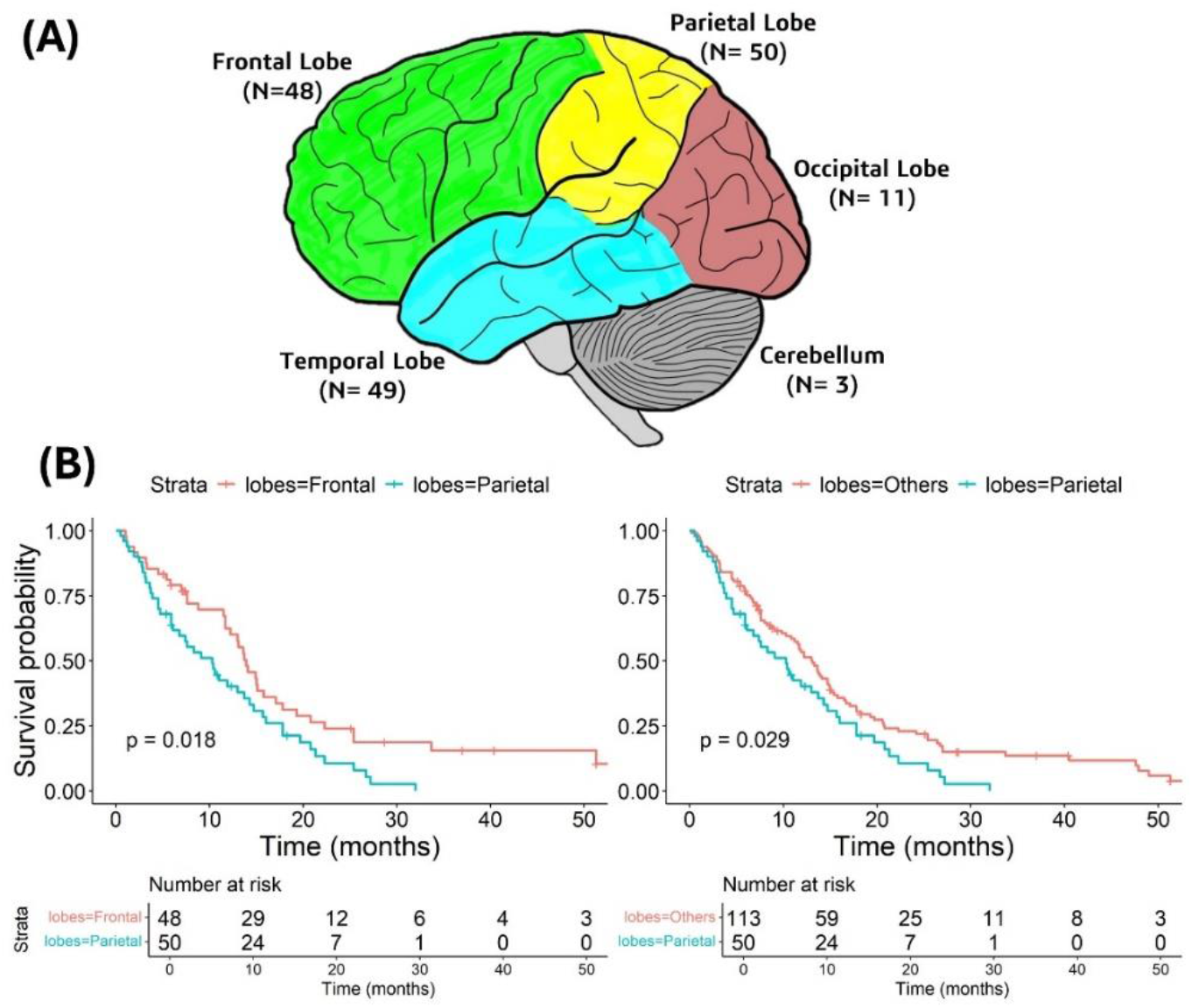
(A) Distribution of GBM Tumor Samples Across various Brain Regions. The sample sizes (N) for each brain region are indicated in parentheses. (B) Kaplan Meier plots showing the survival outcomes of GBM patients from various brain regions.

### 3.2 Tumor Location Impact on Survival Outcome

We conducted multiple comparisons between different tumor locations for their differential survival outcome using Kaplan-Meier statistics as mentioned in method sections. Significant differences in survival outcomes were observed between the parietal and frontal lobe tumors, as well as between parietal and other lobe tumors (P<0.05) **(Figure 1B and Supplementary Figure S1)**. Interestingly, parietal lobe tumors showed poor survival in both comparisons, which warranty a further investigation on genomic landscape of these tumors to identify the genetic basis of their poor survival outcome.

### 3.3 Location Specific Genomic Characteristics

We observed a significant poor survival in patients with parietal tumor location. To identify the genetic differences of parietal lobe tumors with frontal lobe tumors, we conducted a comprehensive comparative genomic analysis.

#### 3.3.1 Somatic Mutation Landscape

We conducted a detailed analysis of the mutational profiles of tumors located in parietal lobe (N=34) and frontal lobe (N=26), to identify differences in their genomic landscape. The most frequently altered genes in the parietal lobe were *PTEN* (35%), *EGFR* (24%), *NF1* (15%), *PCLO* (15%), and *SPTA1* (15%) **(Figure 2A)**. On the other hand, in frontal lobe, the most mutated genes were *EGFR* (27%), *TP53* (23%), *SPTA1* (19%), and *SYNE1* (15%) (**Figure 2B**). The Fisher statistical test results revealed a notable difference in *PTEN* mutations between parietal and frontal lobe tumors **(Supplementary Table 2)**. Specifically, *PTEN* mutations were detected significantly higher in parietal lobe tumors (n=12, P<0.05). Of these 12 parietal lobe tumors, four tumors have missense mutations and two have frameshift deletions in the PTPc domain and four frameshift deletions and two nonsense mutations in the C2 domain **(Figure 2C)**. In contrast, three subjects with frontal lobe tumors exhibited *PTEN* mutations, all of which were missense mutations, with two occurring in the PTPc domain and one in the C2 domain. *PTEN* loss-of-function mutations i.e., nonsense, frameshift and indel variants, are present exclusively in parietal lobe tumors, which could potentially contribute to the poor survival outcomes of parietal lobe tumor subjects [29]. Additionally, frameshift mutations in *PTEN* are known to be associated with resistance to chemotherapeutic drugs [30], which further aggravate the prognosis for these patients. Additionally, we observed that four frontal lobe tumors had *SYNE1* missense mutations (P<0.05) and two samples had mutations in *URGCP* (P<0.05).

**Figure 2.**
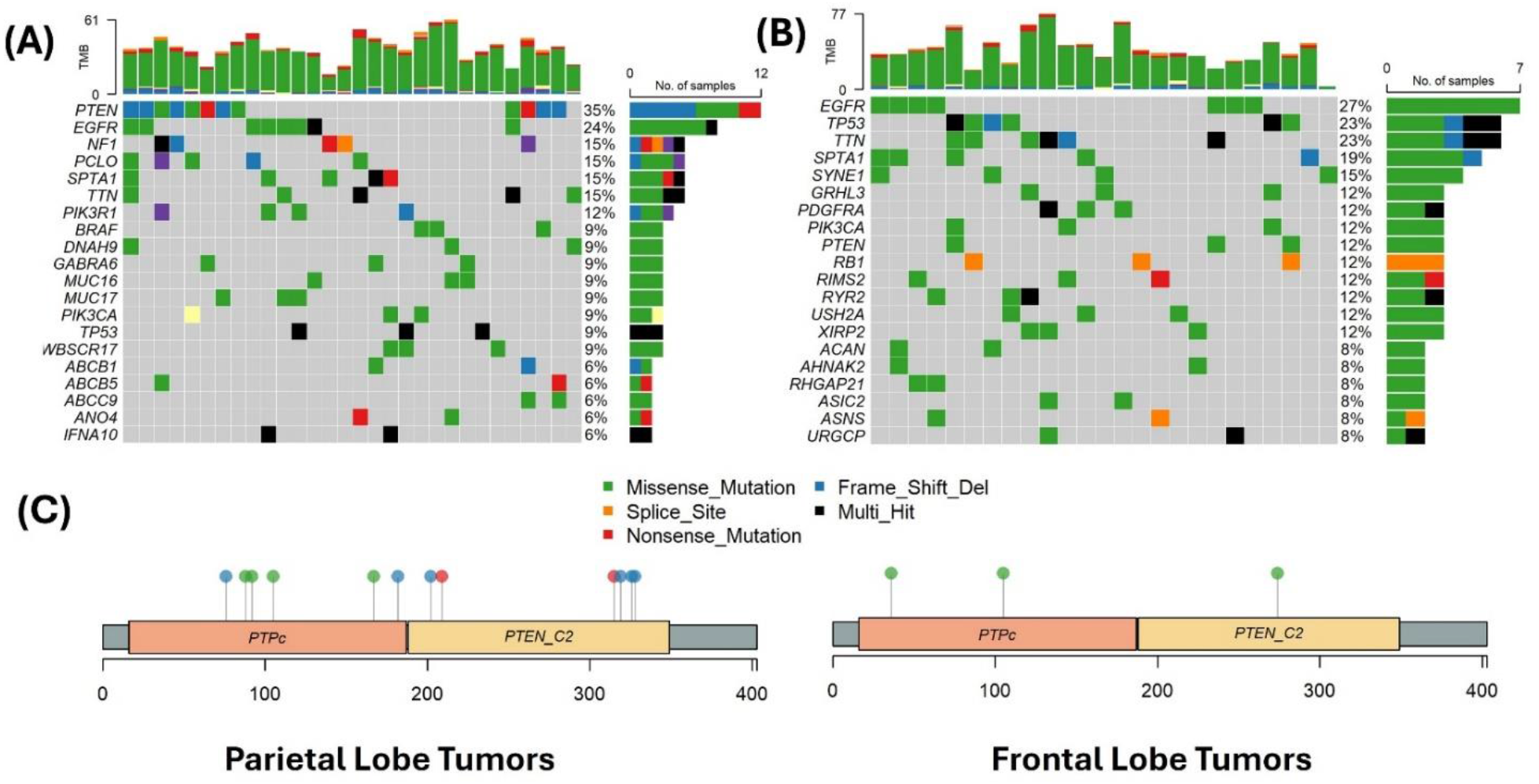
Somatic Mutation Landscape in parietal and frontal lobe tumors (A) Oncoprint of the top mutated genes in the parietal (left) and (B) Frontal (right) lobe tumors. (C) Lollipop plot illustrating the distribution of mutations in the *PTEN* gene across the parietal and frontal lobe tumors. Mutations are marked by colored boxes/circles on the plots, corresponding to different types of mutations.

We also performed mutation co-occurrence and mutual exclusivity analysis to understand genetic interactions. In the Parietal lobe, *IFNA10* mutations significantly co-occurred with *SPTA1* mutations, and *TP53* mutations with *PIK3R1* mutations (P<0.05). Additionally, *BRAF* mutations co-occurred with both *CDH7* and *CD3EAP* mutations, while *CDH7* also co-occurred with *CD3EAP* (P<0.01). In the Frontal lobe, significant co-occurrences were observed between *ASIC2* and *PDGFRA* mutations (P<0.01) (**Figure 3**). These distinct co-occurring mutated genes across the parietal and frontal regions underscore the varying genomic landscapes and the heterogeneity among tumors with different location.

**Figure 3.**
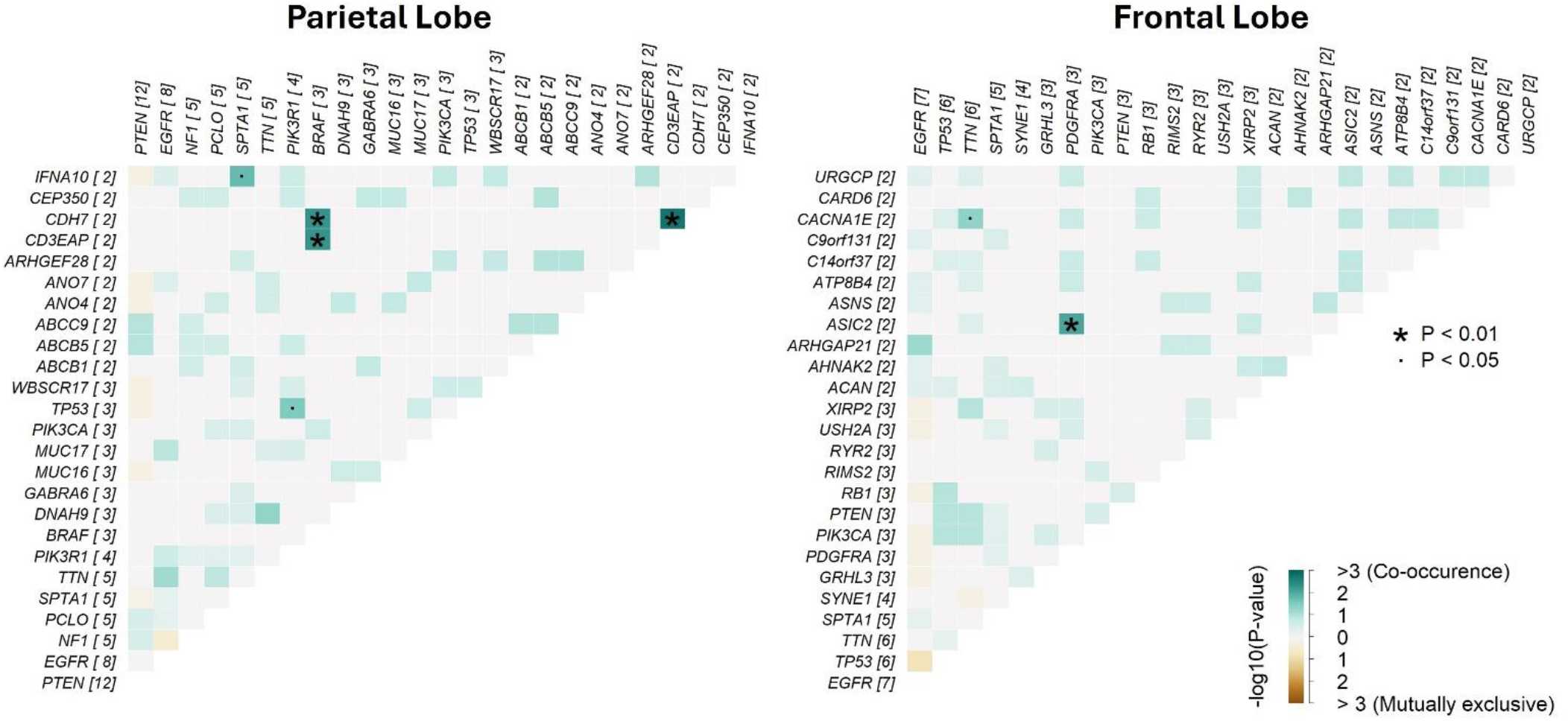
Co-occurrence and mutual exclusivity plot of somatic mutations in parietal and frontal lobe tumors. Asterisks indicate statistically significant interactions.

#### 3.3.2 Fusion Gene Analysis

In our analysis of fusion genes, 34% of parietal lobe tumors were found to harbour 62 fusion genes, while 29% of frontal lobe tumors harboured 40 fusion genes **(Supplementary Table 3)**. Notably, parietal lobe tumors contain two prominent fusion genes, *FGFR3-TACC3* and *EGFR-SEPT14*, which are implicated in driving oncogenesis **(Figure 4)** [31]. No such fusion genes were found in frontal samples.

**Figure 4.**
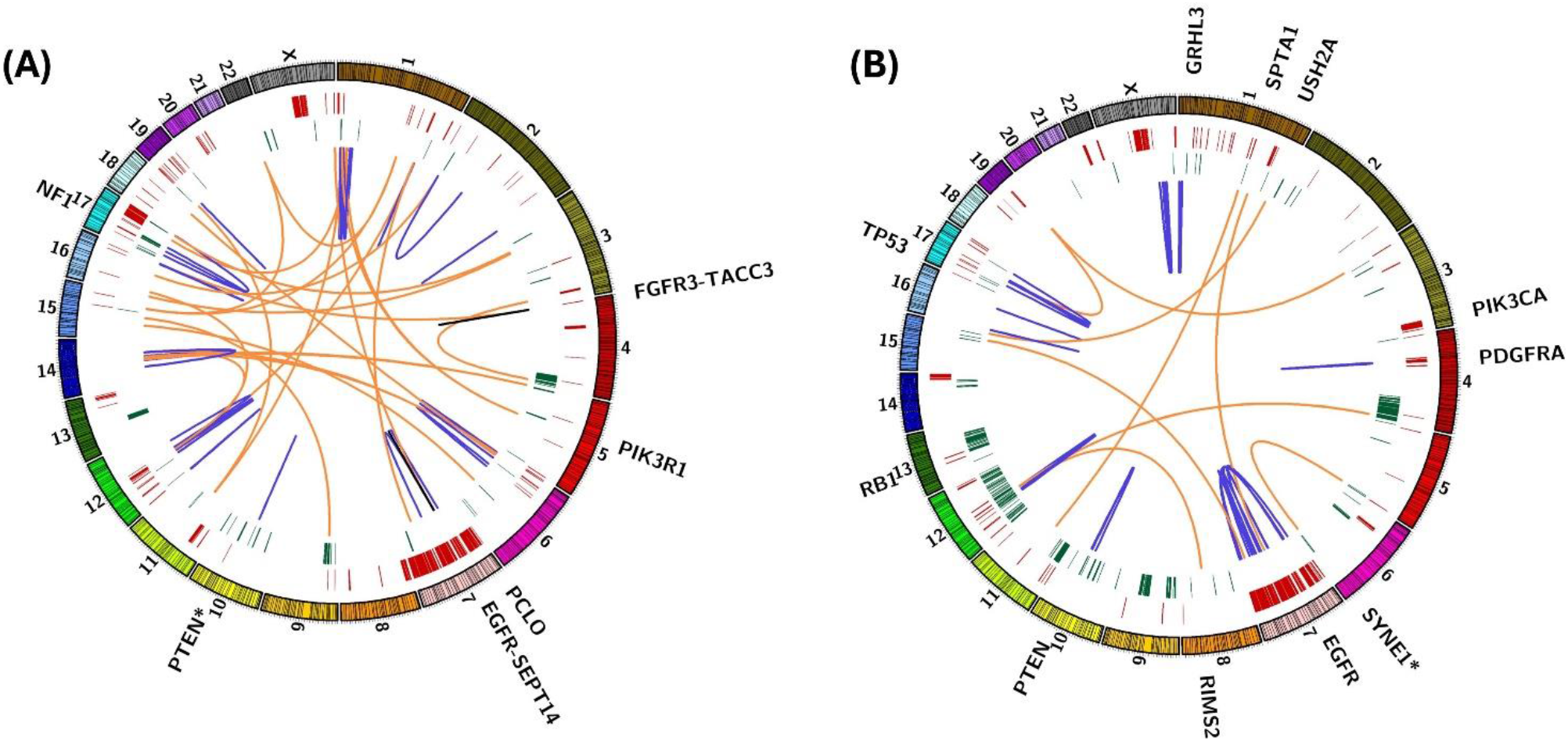
*Circos* plot illustrates the genomic landscape of tumors in the (A) Parietal Lobe (left) and (B) Frontal Lobe (right). The outermost track shows chromosomal locations, with genes labeled outside indicating those with a mutation frequency greater than 10%. Genes labeled with an asterisk (*) are significant based on a Fisher’s test (P<0.05). The next two inner tracks depict copy number alterations, with amplifications (red) and deletions (green) shown from outer to inner. The central links illustrate gene fusions, where blue links represent intrachromosomal interactions, and orange links represent interchromosomal fusion genes. The black link in the parietal lobe indicates the most prominent fusion responsible for tumorigenesis.

Multiple fusion events were detected in a substantial proportion of both parietal and frontal lobe tumors. These fusions occur both inter-chromosomally and intra-chromosomally. Two major genomic hotspots were identified on chromosomes 7 and 12 in both datasets. Specifically, the highest number of fusions in parietal lobe tumors were observed on chromosomes 1 (12/62), 7 (7/62), and 12 (6/62), whereas in frontal lobe tumors, the highest numbers were found on chromosomes 7 (11/40) and 12 (5/40).

#### 3.3.3 Copy Number Variation (CNV) Analysis

Copy number alteration analysis in both datasets revealed significant gene copy number gains and losses that may influence the survival outcome. 31 genes demonstrated significant copy number gains in frontal lobe tumors, while nine genes (two in parietal lobe tumors and seven in frontal lobe tumors) showed significant copy number losses. Among the significantly altered genes listed in **Supplementary Table 4**, *CRYL1* and *SAP18*, located on chromosome 13q12.11, showed copy number gains in 10% (5 out of 47) of frontal lobe tumors. *YES1*, located on chromosome 18p11.32, exhibited copy number loss in 29% (14 out of 47) of frontal lobe tumors. Alterations in these genes play significant roles in tumor progression [32,33]. Transcriptomic comparison of these genes between parietal and frontal lobe tumors revealed significant changes in mRNA levels (P<0.05) **(Supplementary Figure S2)**, with upregulation of *YES1* in parietal lobe tumors is associated with chemotherapeutic drugs resistance [34] and downregulation of *CRYL1* and *SAP18* is correlated with immune-infiltration, EMT [35] and activation of Nf-κB pathway, cell invasion, and angiogenesis respectively [36].

#### 3.3.4 Differential Gene Expression Analysis

In the differential gene expression (DGE) analysis between the parietal lobe (N=19) and frontal lobe (N=15) tumors, 17 genes found to be differentially expressed (P_adj_ < 0.05). Among these, 7 genes are upregulated in the parietal lobe tumors, including *PITX2, SLC9A7, CYP4F3, HOXB13, DTHD1, ATP6VOE2-AS1*, and *EREG*, while 10 genes are downregulated, such as *TNF, PCDHGB5, OR2L13, CXCR2P1, PCDHB6, HHIP, RBP3, KCTD4, MYO22*, and *ABHD12B* **(Figure 5 and Table 1)**. This analysis identified key genes that may contribute to the unique pathological and functional properties of the parietal lobe compared to frontal lobe tumors. These differentially expressed genes may have different impacts on tumor progression and survival outcomes, highlighting the importance of anatomical location in gene expression in glioblastoma.

**Table 1:** Differentially Expressed Genes between patients having tumor in parietal and frontal lobe.

| Gene Symbol | Log2FC | Up/Down | Padj | Gene Description |
| --- | --- | --- | --- | --- |
| **PITX2** | 3.753818 | Up | 0.024584 | Paired Like Homeodomain 2 |
| **HOXB13** | 3.469185 | Up | 0.035627 | Homeobox B13 |
| **EREG** | 3.744343 | Up | 0.037785 | Epiregulin |
| **SLC9A7** | 1.230717 | Up | 0.037785 | Solute Carrier Family 9 Member A7 |
| **CYP4F3** | 2.625896 | Up | 0.043283 | Cytochrome P450 Family 4 Subfamily F Member 3 |
| **DTHD1** | 2.684793 | Up | 0.043283 | Death Domain Containing 1 |
| **ATP6V0E2-AS1** | 0.821265 | Up | 0.044278 | ATP6V0E2 Antisense RNA |
| **PCDHGB5** | -2.80616 | Down | 0.007907 | Protocadherin Gamma Subfamily B, 5 |
| **TNF** | -2.13195 | Down | 0.008486 | Tumor Necrosis Factor |
| **ABHD12B** | -1.55723 | Down | 0.035627 | Abhydrolase Domain Containing 12B |
| **RBP3** | -3.76488 | Down | 0.035627 | Retinol Binding Protein 3 |
| **OR2L13** | -3.42904 | Down | 0.037685 | Olfactory Receptor Family 2 Subfamily L Member 13 |
| **CXCR2P1** | -2.54734 | Down | 0.037785 | C-X-C Motif Chemokine Receptor 2 Pseudogene 1 |
| **HHIP** | -1.71272 | Down | 0.037785 | Hedgehog Interacting Protein |
| **KCTD4** | -2.50478 | Down | 0.037785 | Potassium Channel Tetramerization Domain Containing 4 |
| **MYOZ2** | -2.02546 | Down | 0.037785 | Myozenin 2 |
| **PCDHB6** | -2.04313 | Down | 0.037785 | Protocadherin Beta 6 |

**Figure 5.**
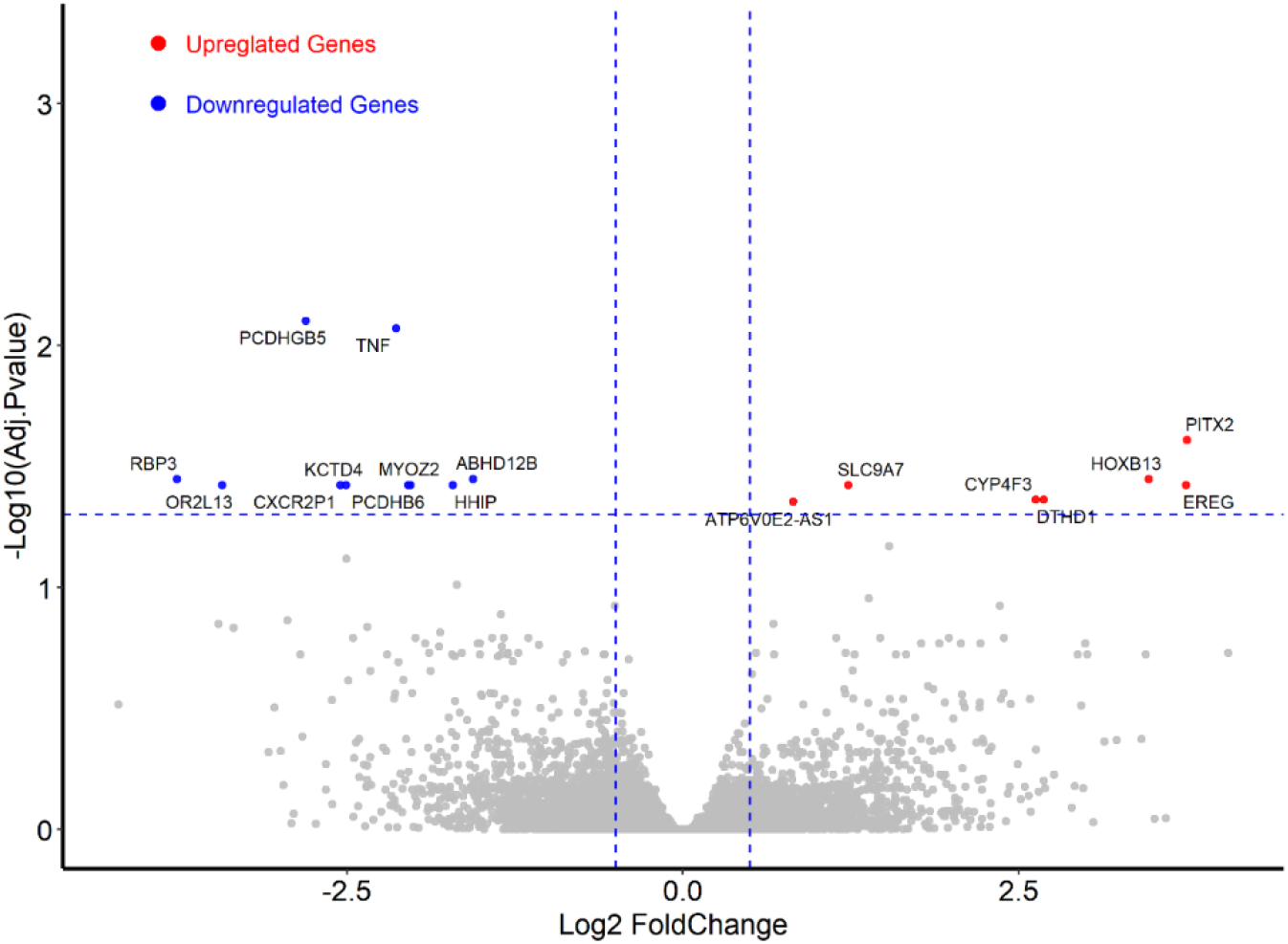
Differentially expressed genes between parietal and frontal lobe tumors. Upregulated genes (log2FC > 0.5) are shown in red, and downregulated genes (log2FC < -0.5) are shown in blue. The significance threshold is set at an Padj < 0.05.

### 3.4 Genomic differences between Parietal and Other Region Tumors

A comparison of mutation rates between tumors in the parietal lobe and those in other brain lobes reveals that *PTEN* mutations occur in 35% of parietal lobe tumors compared to 20% in other lobes; *EGFR* mutations are observed in 24% versus 29%; *NF1* mutations in 15% versus 8%; *SPTA1* mutations in 15% versus 14%; and *TP53* mutations in 9% versus 15%, respectively **(Supplementary Figure S3a)**. However, none of these gene mutations were found to be statistically significant in Fisher’s exact test.

Significant co-occurrences were observed between *RYR2* and *RPL5*, as well as *OR5M3* (P<0.01). Similarly, *HMCN1* co-occurs with *TP53*, and *RPL5* co-occurs with *OR5M3* (P<0.05). These co-occurring mutations may contribute to the better survival rates observed in subjects with tumors in other lobes. Notably, these co-occurring mutations were absent in parietal lobe tumors, which are associated with poorer survival outcomes **(Supplementary Figure S3b)**.

Our fusion genes analysis revealed that 34 samples from other lobes harbour a total of 117 fusions. Among these, 23 samples exhibit multiple fusions occurring both inter-chromosomally and intra-chromosomally. We identified three major genomic hotspots located on chromosomes 1, 7, and 12. Importantly, no oncogenic fusion genes were found in these samples. In contrast, parietal lobe tumors exhibited distinct genomic alterations. Four genes showed significant copy number gains, while one gene was deleted specifically in the parietal lobe. Notably, the tumor suppressor gene *LINC00290*, located on 4q34.3, was deleted in 6% (3 out of 49) of parietal lobe tumors [37,38]. The loss of *LINC00290* may contribute to tumor progression and is potentially linked to the poorer survival outcomes observed in subjects with parietal lobe tumors.

In transcriptomics comparison, 15 genes showed significant differential expression (P_adj_ < 0.05). 13 genes being upregulated in the parietal lobe, including *ESR2, LGR6, HS3ST5, SLC7A10, IGFBP6, SLC4A11, ARHGEF35, HS3ST3, CNTNAP3, PAPPA, CHST1, EFCA4BL*, and *IGFBP6*. Meanwhile, two genes, *ALOX15* and *ZNF560*, are downregulated **(Supplementary Figure S4 and Supplementary Table 5)**.

### 3.5 Radiomics Analysis between Parietal and Frontal Lobe tumors

In the radiomic analysis comparing parietal and frontal lobe tumors, 1213 radiomic features were extracted from T1ce scans of each subject. These features include 17 shape features, 18 first-order statistics, 74 texture features, 368 Laplacian of Gaussian (LoG) features, and 736 wavelet features **(Supplementary Figure S5)**. The features were normalized using the “scale” function in R (v4.2.1) [23]. A comparative analysis identified nine significant radiomic features distinguishing parietal from frontal lobe tumors, based on the Wilcoxon test (p < 0.05). Of these, eight were wavelet features and one was a texture feature, spanning various sub-bands such as HLL, LLH, LLL, and HLH **(Supplementary Table 6)**.

A multivariate Cox proportional hazards model was used to assess the relationship between these radiomic features and patient survival in parietal tumors, revealing three features significantly associated with survival outcomes. Low *LLL_GLDM_DependanceEntropy* (HR = 2.47, p = 0.014) and high *HLL_FirstOrder_Mean* (HR = 2.90, p = 0.025) were associated with increased risk and poorer survival, while high *HLL_GLCM_ClusterShade* (HR = 0.16, p = 0.043) was linked to better survival **(Table 2)**. These findings suggest that higher signal intensities and more uniform textures correlate with worse prognosis and poor survival outcome. Conversely, greater asymmetry (indicated by high ClusterShade) is associated with lower risk. The forest plot visualization (**Figure 6**) of the Cox model results illustrates the hazard ratios and confidence intervals for these features, reinforcing their prognostic significance for parietal tumors.

**Table 2:** Significant radiomic features that distinguish between tumors in the parietal and frontal lobes, along with the results of the Cox proportional hazard model regression within the parietal lobe. *Statistically significant features (P < 0.05) are indicated, with red color representing high risk and blue representing low risk.

| Radiomic Features | Comparative Analysis | Multi-variate Cox Hazard Analysis |  |  |  |
| --- | --- | --- | --- | --- | --- |
|  | p-value | beta | HR | CI | p-value |
| *LLL_GLDM_DependenceEntropy* | 0.01* | 0.91 | 2.47 | 1.2 – 1.2 | 0.01* |
| *HLL_FirstOrder_Mean* | 0.01* | 1.06 | 2.90 | 1.14 – 7.38 | 0.03* |
| *HLL_GLCM_ClusterShade* | 0.03* | -1.81 | 0.16 | 0.03 – 0.95 | 0.04* |
| *HLH_GLCM_ClusterShade* | 0.03* | 2.26 | 9.61 | 0.84 – 109.76 | 0.07 |
| *LLL_GLCM_SumEntropy* | 0.04* | -2.28 | 0.10 | 0.002 – 3.98 | 0.22 |
| *GLCM_SumEntropy* | 0.04* | 1.21 | 3.37 | 0.11 – 101.66 | 0.48 |
| *LLL_GLDM_LDHGLE* | 0.04* | 0.20 | 1.22 | 0.65 – 2.3 | 0.53 |
| *LLL_GLCM_Correlation* | 0.04* | -0.16 | 0.85 | 0.47 – 1.53 | 0.58 |
| *HLH_FirstOrder_Mean* | 0.04* | -0.15 | 0.86 | 0.34 – 2.17 | 0.75 |

*LDHGLE: LargeDependenceHighGrayLevelEmphasis; HR: Hazard Ratio; CI: Confidence Interval*

**Figure 6.**
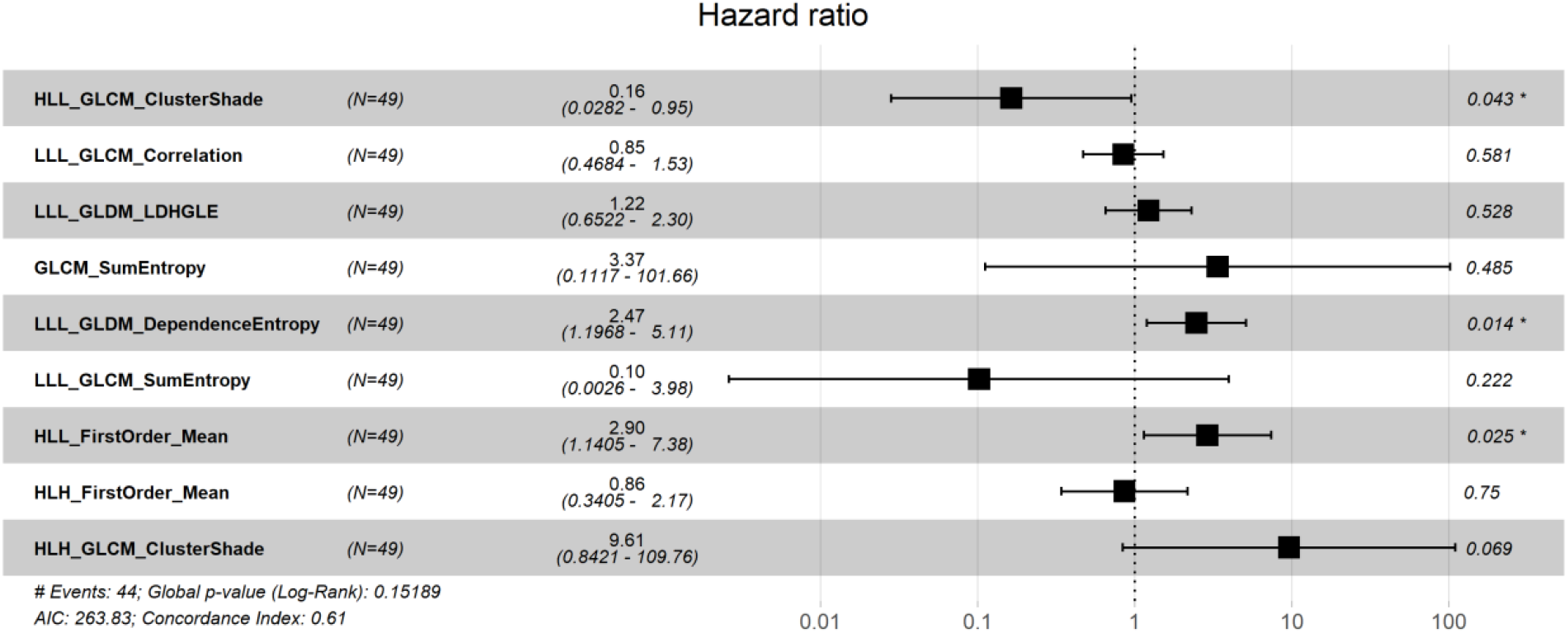
Forest plot of cox proportional hazards multivariable regression on overall survival in parietal lobe tumors (N=49) showing hazard ratios (HR) and 95% confidence intervals (CI) for nine radiomic features. Significant features (P < 0.05) are marked with asterisks (*).

## 4 Discussion

In this study, we investigated the impact of tumor location on survival outcome in glioblastoma, followed by a comprehensive genomic analysis to uncover the genetic aberrations contributing to the differential survival outcomes. To achieve this, we first identified tumor locations using the MNI atlas within the FSL tool. Our survival analysis revealed that GBM patients with tumors in the parietal lobe had poorer survival outcomes compared to those with tumors in other brain region. Notably, there was a significant difference in survival when comparing patients with tumors in the parietal lobe to those with tumors in the frontal lobe. To understand the genetic basis of poor survival outcome in parietal lobe tumor, we conducted a comprehensive genomic analysis, including mutational profiling, fusion genes, copy number variations, and differentially expressed genes (DEGs). We found that *PTEN* in parietal lobe, whereas *SYNE1* and *URGCP* are the most significantly mutated gene in frontal lobe. *PTEN* mutations, particularly consist of loss-of-function mutations i.e., frameshift and non-sense mutations in its C2 domain. Interestingly, *PTEN* loss-of-function mutation known to contribute to chemotherapeutic resistance, potentially leading to worse clinical outcomes for patients with parietal lobe tumors [29,39].

We identified two fusion genes, *FGFR3-TACC3* and *EGFR-SEPT14*, that were exclusively found in tumors located in the parietal lobe. Both fusion genes have been associated with promoting glioblastoma tumorigenesis and may contribute to the poor survival outcomes observed in these tumors. *FGFR3-TACC*3 fusion is reported as oncogenic fusion disrupting normal *FGFR3* signalling in astrocytic differentiation, leading to tumor cells formation [31]. *EGFR-SEPT14* fusion leads to the activation of the *STAT3* signalling pathway allowing cells to grow and divide without the need for external growth signals and promote oncogenesis [40].

In our study, we identified a copy number gain of 13q (*CRYL1* and *SAP18*), a loss in 18p (*YES1*), and a deletion in 4q (*LINC00290*). *CRYL1* (Crystallin lambda 1) and *SAP18* (Sin3 associated protein 18) have copy number gains in frontal lobe tumors, leading to their upregulated expression, which is associated with improved patient survival [35,36]. *YES1* (YES proto-oncogene1) plays a pivotal role in promoting cell proliferation, tumor survival, and invasiveness during tumorigenesis. Loss of proto-oncogene (in frontal lobe tumors) reduces oncogenic processes and improve patient survival. Increased expression of YES1 in parietal lobe tumors is linked to resistance to chemotherapeutics and tyrosine kinase inhibitors in various human cancers and is associated with lower survival rates. [34]. *LINC00290* is a long non-coding RNA recently discovered as a new tumor suppressor gene, frequently showing homozygous deletion in various cancers [37,38]. In our study, this non-coding gene was explicitly deleted in parietal lobe tumors, potentially contributing to poor survival in these patients.

Our differential gene expression (DGE) analysis between parietal and frontal lobe tumors revealed seven genes that were significantly upregulated, and ten genes were significantly downregulated in parietal lobe tumors. Interestingly, all upregulated genes are well known to play crucial roles in promoting tumor aggression in different cancer types. *PITX2* and *HOXB13* are transcription factors that regulate pathways involved in cell growth, migration, and invasion, which are critical processes in tumor progression. *PITX2* is also linked to the activation of the WNT/β-catenin and TGF-β signaling pathways [41,42], while *HOXB13* enhances tumor proliferation through its regulation of transcriptional complexes [43].

Other upregulated gene, *DTHD1* is associated with the regulation of mRNA modifications which contribute to enhanced tumor aggressiveness [44,45]. *SLC9A7* (*NHE7*) is involved in organellar homeostasis and vesicular trafficking, processes that are important for the survival and growth of tumor cells [46]. Another gene, *LGR6*, is significantly overexpressed in parietal lobe tumors compared to other lobe tumors and has been implicated in the activation of the WNT signaling pathway, which is a well-known driver of tumor formation [47]. Lower expression levels of *ALOX15* linked to an increased risk of disease relapse [48]. This suggests that parietal lobe tumors with reduced *ALOX15* expression are more susceptible to cancer relapse, thereby decreasing the likelihood of survival. These molecular alterations indicate that tumors in the parietal lobe may have unique genomic signatures contributing to their aggressive behaviours and poor prognosis. These findings highlight the importance of considering molecular heterogeneity when developing targeted therapeutic interventions for patients with tumors in different brain lobes.

Radiomics is rapidly emerging as a transformative field in modern radiology. These non-invasive digital fingerprints enable early diagnosis and open doors to safer, more personalized treatments by identifying key tumor characteristics and their associated genomic factors [14]. As reported higher rad-scores have been linked to C5aR1 expression [49] and tumor-infiltrating macrophages [50], while *GLDM_DependenceEntropy* is correlated with recurrence and metastasis [51]. Identifying such radiomic features enhances treatment planning, allowing clinicians to tailor therapies based on a tumor’s unique radiomic and molecular profile. In our analysis, 1213 radiomic features per sample encompassing shape, first-order statistics, texture, LoG, and wavelet features were examined. We identified nine significant features that distinguish parietal from frontal lobe tumors, with eight being wavelet-based and one texture-based. These features capture critical aspects such as intensity variation, entropy, and pixel dependency. Through a Cox proportional hazards model, we found that lower Dependence Entropy and higher First-Order Mean were associated with poorer survival outcomes, while higher ClusterShade was linked to improved survival, emphasizing the prognostic importance of tumor texture and intensity variations.

Our study warrants a further research and experimental validation of these genomic aberrations for their roles in treatment responses in GBM patients and survival outcome. Investigating these genomic aberrations can further lead to the development of targeted therapies and personalized medicine. Understanding how these genomic alterations influence GBM progression, specifically in parietal lobe tumors, and treatment response could significantly impact the management and outcomes of GBM patients. Our findings also underscore the potential of radiomic features as non-invasive biomarkers for predicting tumor behaviour and survival outcomes, offering valuable insights for personalized treatment strategies. However, limitations such as the availability of datasets, particularly both genomic and MRI data, highlight the need for more extensive studies. More comprehensive data collection, including pre- and post-imaging data will further increase the impact of these findings.

## 5 Conclusion

Our comprehensive analysis of glioblastoma patients revealed that tumors located in the parietal lobe are associated with poor survival outcome compared to the other brain regions. Key genetic alterations, such as the *PTEN* mutation, overexpression of *PITX2, HOXB13, DTDH1*, downregulation of *ALOX15*, along with the presence of *FGFR3-TACC3* and *EGFR-SEPT14* fusion genes, structural alteration in *LINC00290* significantly contribute to the aggressive nature of parietal lobe glioblastomas. Additionally, two radiomic features, lower *LLL_GLDM_DependanceEntropy* and higher *HLL_FirstOrder_Mean* shows significant correlation with increased risk and poor survival outcome. These findings highlight the potential for targeted therapies and personalized treatment approaches based on tumor location, genetic profile and non-invasive biomarkers. However, the limited availability of genomic datasets compared to MRI data underscores the need for large cohort studies to better understand the molecular mechanisms driving glioblastoma progression and treatment resistance and their impact on patient survival outcome.

## 6 Data Availability

The Genomics dataset *(gbm_tcga_pub2013)* analysed in this study is available in cBioPortal: *www.cbioportal.org* and imaging data *(TCGA-GBM)* is available at The Cancer Image Archive database (TCIA): *https://www.cancerimagingarchive.net/*.

## 7 Author Contribution

Kavita Kundal and Rahul Kumar conceived and designed the study. Kavita Kundal conducted the data collection, data analysis and manuscript writing under the supervision of Rahul Kumar. All authors read and approved the final version of the manuscript.

## 8 Acknowledgments

The authors acknowledge The Cancer Imaging Archive (TCIA) for providing access to the imaging data used in this study. They also acknowledge the infrastructure support provided by the Indian Institute of Technology Hyderabad. KK acknowledges the financial support from the Ministry of Education (MoE), India.

## 9 Ethics approval and consent to participate

Ethics approval and consent to participate: Databases such as TCGA and TCIA are publicly accessible, and the patients included in these databases have obtained the necessary ethical approvals. Researchers can freely download and use the data for research purposes and publish related findings. The data used in our study were obtained under a license from TCIA. As our study is based on open-source data, there are no ethical concerns or conflicts of interest.

## 10 Consent for publication

Not applicable.

