## Supplementary File 1 for "Unravelling the Impact of Tumor Location on Patient Survival in Glioblastoma: A Genomics and Radiomics Approach"

### Supplementary Figures

**Figure S1:** Kaplan Meier plots of Glioblastoma Patients by Brain Region.

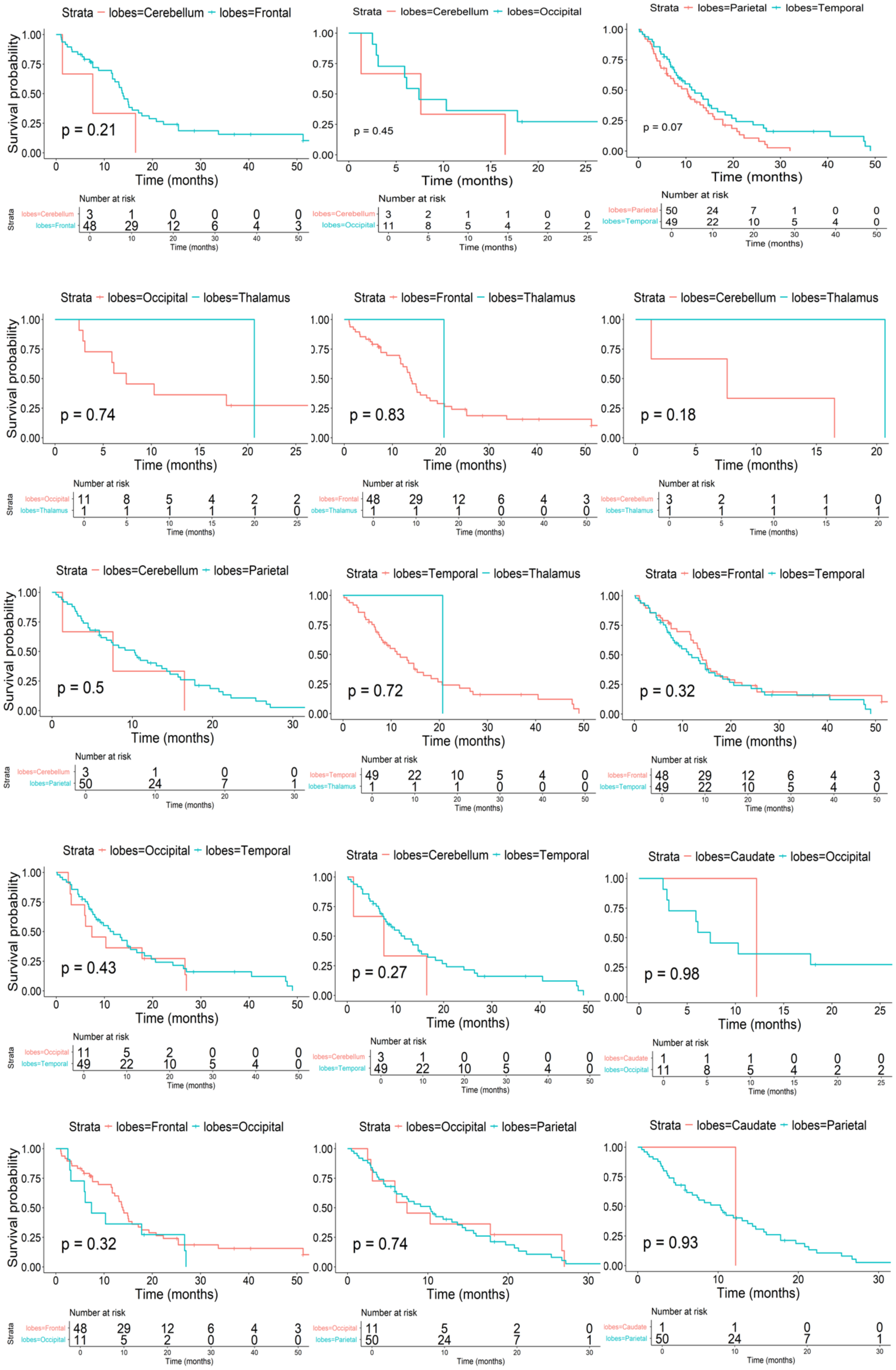

**Figure S2:** Boxplots showing normalized expression levels in the Frontal and Parietal lobe tumors. *CYRL1* (copy number gain), *SAP18* (copy number gain), and *YES1* (copy number loss) show significant differences P<0.05.

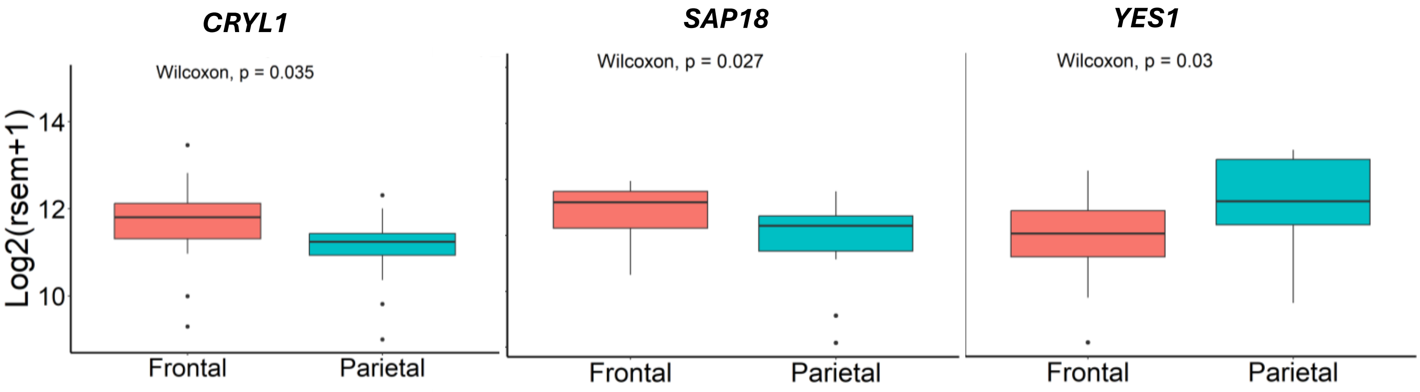


**Figure S3:** (A) Co-bar plot showing mutation frequencies in parietal lobe tumors (left) compared to tumors from other brain lobes (right). (B) Co-occurrence and mutual exclusivity plot of mutations across tumors from other brain lobes. The plot highlights significant co-occurrence and mutual exclusivity patterns. Asterisks indicate statistically significant interactions.

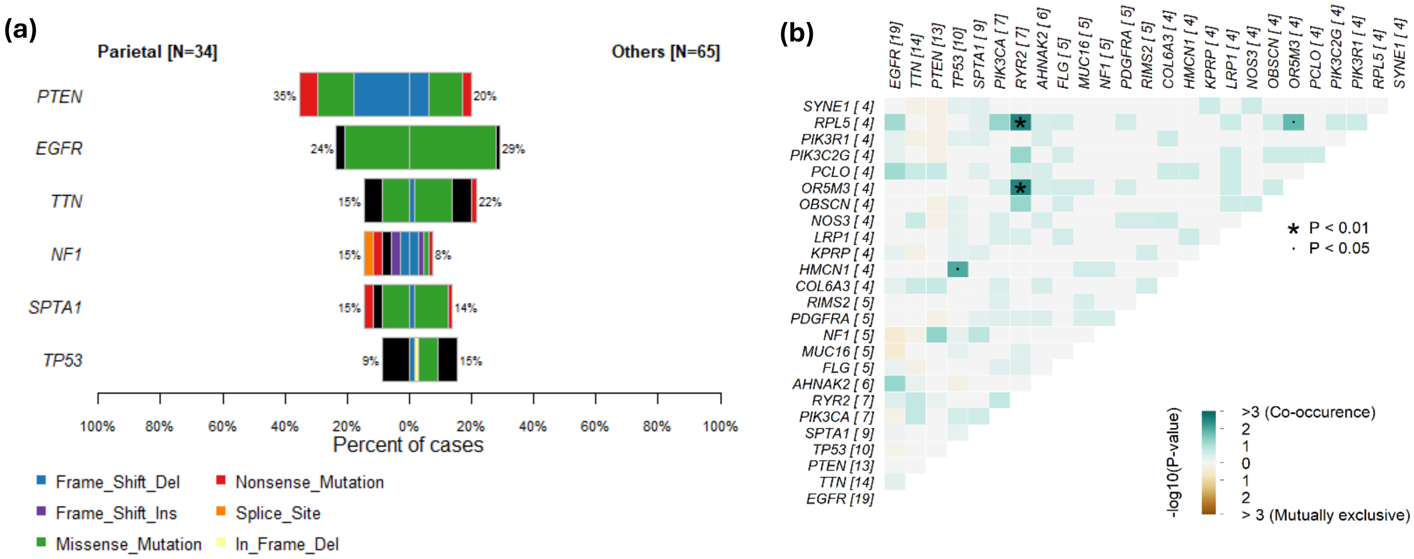

**Figure S4:** Differentially expressed genes between parietal lobe versus other brain region samples. Upregulated genes (log2FC > 0.5) are shown in red, and downregulated genes (log2FC < -0.5) are shown in blue. The significance threshold is set at an Padj < 0.05.

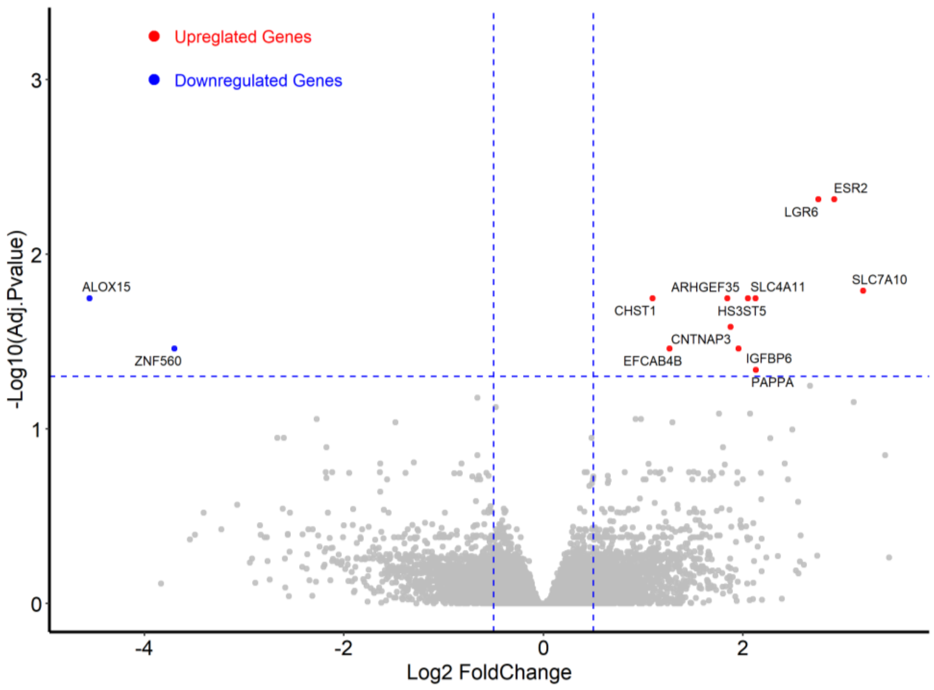


**Figure S5: :** Heatmap displaying 1213 radiomic features clustered based on subjects with tumors in the frontal and parietal lobes. Features are color-coded by 5 categories (FirstOrder, LoG, Shape, Texture, Wavelet), and top annotation differentiates brain tumor location, with pink representing frontal lobe tumors and purple representing parietal lobe tumors

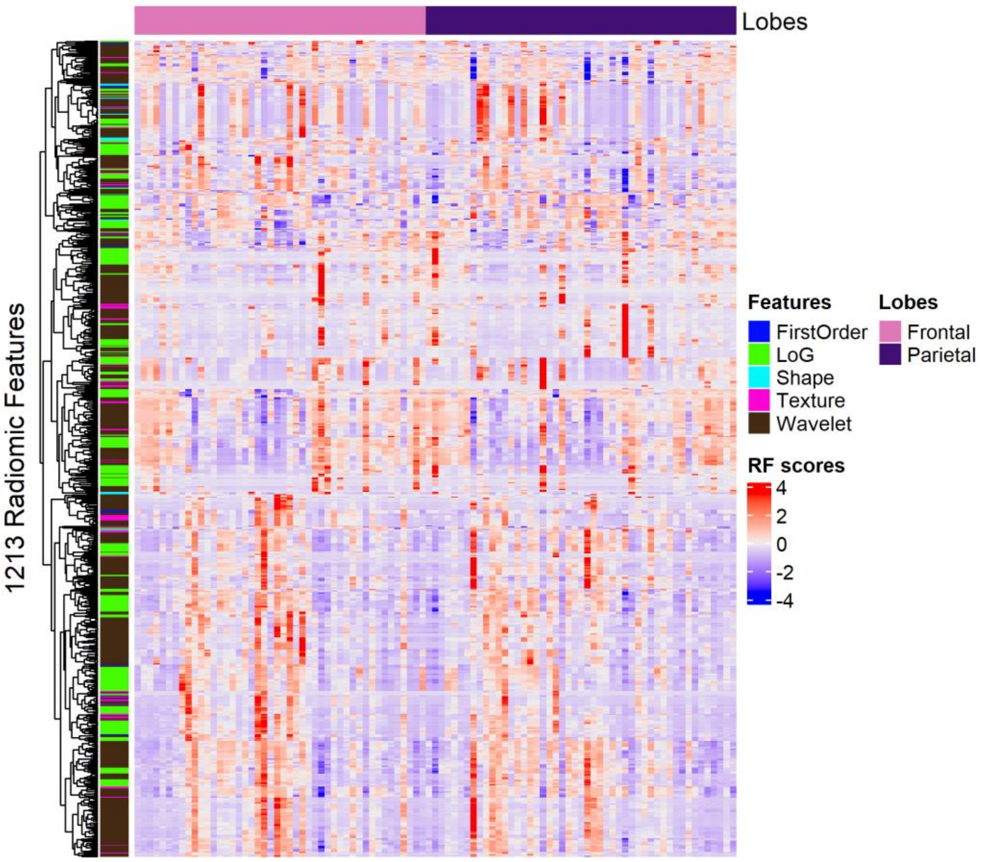
